# Covid-19 Vaccination in India: An Exploratory Analysis

**DOI:** 10.1101/2023.02.01.23285134

**Authors:** Sandip K. Agarwal, Maharnab Naha

**Affiliations:** IISER Bhopal

## Abstract

Our study is designed to explore the patterns in covid vaccination coverage in India at the district level. We use data from the first six months of covid vaccination drive in India that we combine with several other administrative data to create a unique data set that facilitates heterogeneity analysis across different vaccination phases and districts. We find evidence of past reported infection rates positively correlated with higher first dose covid vaccination outcomes. Higher Deaths as a proportion of district population is associated with lower vaccination uptake but as a percentage of reported infection was positively correlated with first dose covid vaccination. Districts that on average had higher population burden per health centre also had lower covid vaccination rates. Vaccination rates were lower in rural areas relative to urban areas whereas the association with literacy rate was positive. A higher vaccination rate among the population with higher blood pressure and hypertension (one of the comorbidities with covid infection) was observed while vaccination rates were lower among pregnant women and breastfeeding mothers. Districts with higher percentage of children with complete immunisation were associated with higher covid vaccination rates whereas low vaccination rates were observed in districts that reported relatively higher percentage of wasted children.

## Introduction

The adverse impacts of COVID-19 pandemic has been felt on all spheres of human life with unprecedented challenges to public health and the economy. Herd immunity has been the key to minimise disruptions caused due to the pandemic. Sooner the population acquires herd immunity, sooner human life and activity would restore to pre-pandemic levels [1, 2]. However, relying upon the natural process of building up herd immunity would have been slow that would have given room to a prolonged pandemic. For governments and policy makers around the world, mass inoculation against COVID-19 has been the inevitable response to tackle the pandemic [3, 4].

While scientists have developed vaccines for COVID-19 at unprecedented speed, the process of translating this discovery into a manufactured product on a large scale and making it available to the masses through an uninterrupted and efficient supply chain is an inevitable component of mass inoculation [5, 6]. Vaccine shortages due to limited manufacturing capacity or due to inefficiencies in the supply chain would adversely affect the vaccination drive. In addition, a successful and an effective vaccination campaign requires much more than the availability of a safe and effective vaccine. Availability of a vaccine does not mean that people will rush to get inoculated. Introduction of a new vaccine demands rigorous research surrounding psychological, social and political aspects to assess public trust in the vaccine as much as it demands scientific rigorous evidence on safety and efficacy of the vaccine [7, 8]. Therefore, the observed vaccination outcomes are a result of interaction between supply and demand for the vaccine - where either of inadequate demand or supply or both would lead to low vaccination rates.

In the context of vaccine demand, a delay in acceptance or refusal of vaccination despite availability of vaccination services has been defined as vaccine hesitancy [9]. In 2019, the World Health Organization (WHO) declared vaccine hesitancy as one of the top ten threats to public health. If vaccine hesitancy is timely addressed, this can avert an adverse public health outcome [10].

COVID-19 vaccination rates had been one of the lowest among the low-and-middle-income-countries (LMICs) [11, 12, 13]. As most LMICs relied upon other vaccine producing nations for the supply of COVID-19 vaccines, vaccination against COVID-19 had been slow due to inadequate supply of vaccines [14, 15]. In addition, a host of complicated demand driven issues surrounding vaccine hesitancy could have further contributed to low vaccination rates as evident from vaccine hesitancy or uptake surveys [16, 11, 17, 18, 19].

Among the LMICs, India is one of the few that produced covid vaccines domestically [20]. In the past decade, India has stood out to be one of the largest vaccine producers in the world with a share of around 60% of vaccine supplies to UNICEF [21, 22, 23, 24, 25, 26]. Covid vaccination campaign in India was one of the largest in the world [27, 28]. In comparison to other LMICs, the proportion of the Indian population vaccinated with covid vaccine along with the pace at which they were vaccinated had been phenomenal [29, 30, 31, 32]. In this article, we analyse the observed covid vaccination rates in India. Our research is built upon a novel data set that was created by combining several administrative data sets and COVID-19 data sets for India. While researchers have analysed the the district level covid infection and fatalities [33, 34], as far as we know, our study is the first one to provide a comprehensive analysis of covid vaccination rates at the district level across the various phases of covid vaccination and the various demographic, socio-economic and health factors associated with the observed vaccination rates. Most existing research studies that have analysed the regional variation in the observed covid vaccination rates in India, research has been mostly limited to the heterogeneity in vaccine rates at the state level or a particular state or district and for early phases of vaccination [35, 36, 37, 38]. Our research makes important contributions to the field of public health research by improving our understanding of factors that are associated with covid vaccination rates in India and in providing directions for future research. Our findings from the Indian covid vaccination drive can significantly contribute to learning of other LMICs regarding the vaccination programs [39].

Our article is organised as the following. Section 2 lays down the background for vaccination in India in general followed by specific details related to covid vaccination campaign in India including the timeline for covid vaccination. Section 3 describes the data and the methodology. Section 4 presents the result and discusses the findings with an attempt to link it with possible reasons that could have led to the findings. Section 5 concludes.

## Background

India had set up the immunisation programme in the 1970s (later renamed as the Universal Immunization Program (UIP)) with the objective of providing life-saving vaccinations that would play a critical role in lowering India ‘s children and neonatal mortality and morbidity rates. While, during the first decade of the UIP ‘s operation, various immunisation coverage levels in India had reached 70-85%, there had been a decline in status since then [40].

Anti-vaccine sentiments in India had never been an organised anti-vaccine movement unlike the west. Prior to the UIP during British India and after independence few prominent Indian leaders who had mass appeal among the people were apprehensive of safety and efficacy of the vaccines or questioned the compatibility of vaccines with their religious beliefs. The UIP met with criticism from a section of elitists on account of being a relatively costly program for a poor country like India [41]. However, since its inception UIP has led to successfully eliminating polio and maternal and neonatal tetanus from India by 2015 [42]. There had been only few lone episodes of resistance to particular vaccines in relatively smaller pockets in India that caught international attention [43, 44].

Covid vaccination drive in India began on January 16, 2021 with two double-dose approved vaccines - Covidshield and Covax. Covid vaccination was voluntary, providing room for delays in accepting the vaccine or refusing to get vaccinated in spite of the availability of a vaccine. Covid Vaccination drive in India was rolled out in phases that prioritised the section of population to get vaccinated first who were at the highest risk of infection [45]. The timeline for the same has been depicted in figure 1.

**Figure 1:**
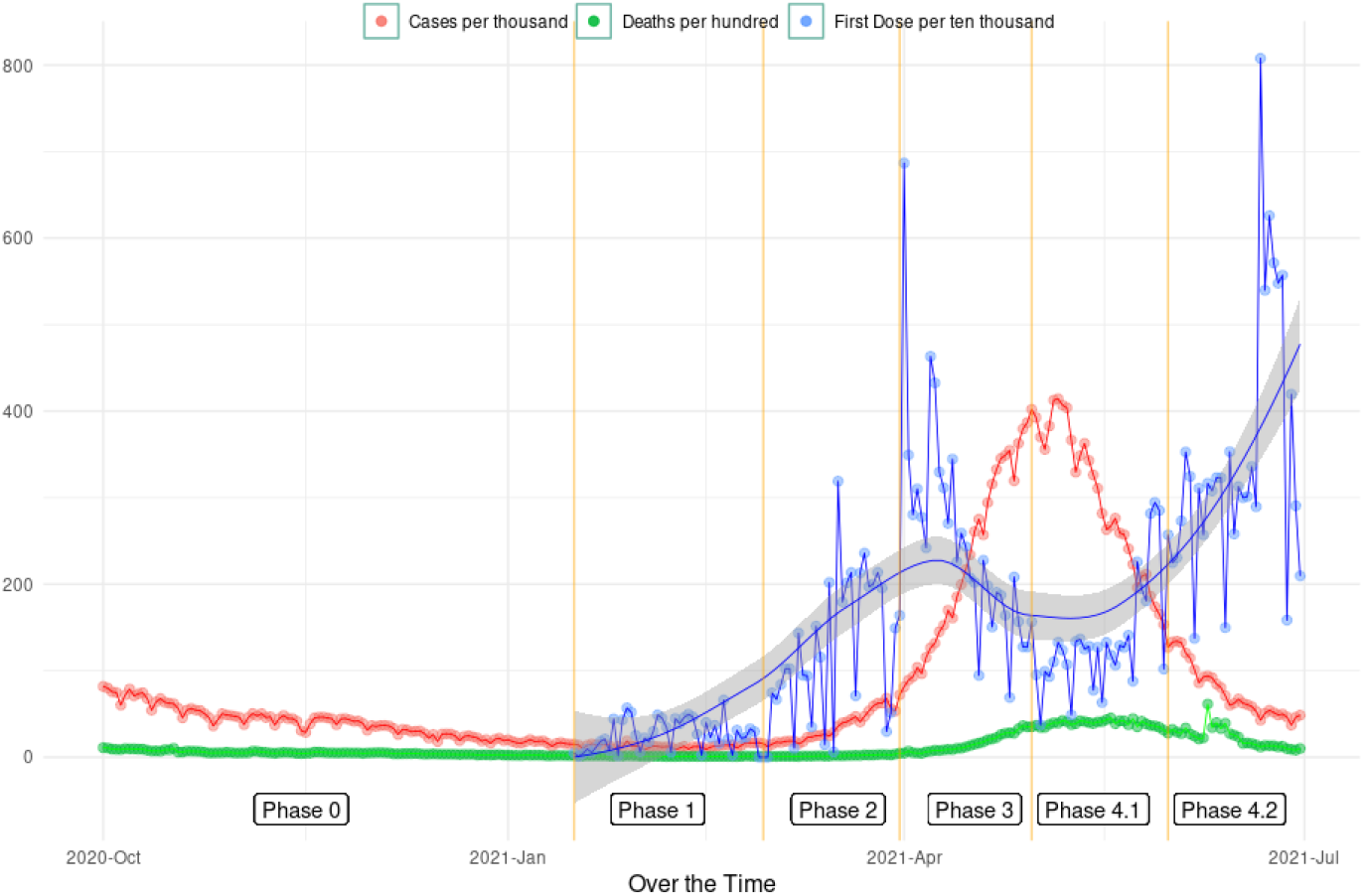
Reported infection rate, Death rate and First dose covid vaccination in India across different phases.

Phase-1 between January 16 to February 28 covered vaccination of all frontline workers, who were actively involved in containing the spread of the pandemic. The frontline workers included health workers, security staff and people who provided essential services to the general public. During Phase-2 in March 2021, the population aged 60 years and above were eligible for vaccination. In addition, individuals aged 45 years and above with comorbidities like hypertension, diabetes, HIV infection etc. too were made eligible for vaccination during phase 2. Phase-3 in the month of April extended COVID-19 vaccination to all individuals aged 45 years and above.The last and the final phase-4 of the vaccination starting May 1 made all adults 18 years of age and above eligible for vaccination.

## Data and Methodology

We used data from various secondary sources to create a unique data set at the district level that allowed us to investigate the trends in vaccination rates across different districts and vaccination phases. In addition, we also included several demographic and health indicator variables that were capable of explaining the variation in the observed vaccination rates.

We obtained district level data for daily covid vaccination numbers as well as daily deaths and daily reported caseload numbers from the Data Development Lab [46]. The daily district level data on vaccination numbers, caseloads and deaths were aggregated across the vaccination phases to facilitate a phase wise analysis of covid vaccination rates. The phase wise data on vaccination numbers, caseloads and deaths were further normalised with district level population to arrive at respective rates. Figure 2 plots the reported infection and fatalities for the entire country over time. The population data used for normalisation was sourced in from the Harvard population database (Dataverse) [47] which provides the latest approximation for all districts in India in 2020.

**Figure 2:**
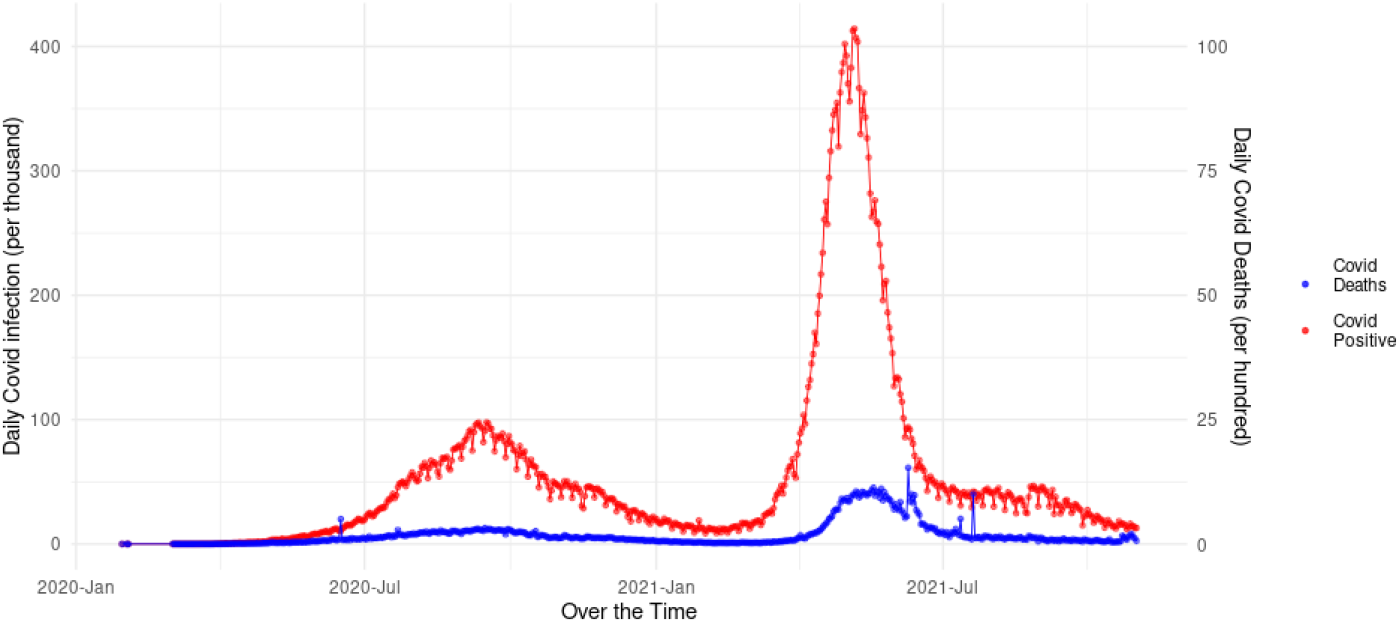
Reported covid infection rates and covid deaths in India over time

As a measure of district level health infrastructure, we used the data from the Rural Health Statistics (RHS) for the year 2020-21 from the Health Information Management System (HMIS) portal of India. The data consists of the number of sub health centres, primary health centres and community health centres in every district. We converted the primary and community health centres into equivalent units of sub-health centres in the ratio 1:6 and 1:24 to arrive at a comparable number of health centres in all districts ([48]). Using the total equivalent number of sub health centres, two health infrastructure variables were constructed. The first one measured the average burden on a sub-health centre in a district, which was computed by dividing the population in that district by the number of equivalent sub health centres in a district. The other infrastructure variable is a measure of distance captured by the infrastructure density, which is constructed by dividing the number of equivalent sub-health centres by geographical area of the district (i.e. number of sub-health centres per square kms). Higher the density in a district, it is likely that smaller would be the average distance travelled to access the health centre.

In addition to health infrastructure, we used several district level health and education indicators from the latest round of the National Family and Health Survey (NFHS) data, which was the fifth round that was collected between 2019-21. NFHS data is an important database that tracks health status over time and administrative units in India that has been collected every four to five years since 1992-93. Some of the notable variables from NFHS-5 that we included in our analysis includes child and maternal health and nutrition, years of schooling, family planning etc.

In the beginning of 2018, Govt. of India identified 117 most backward districts (and called them aspirational districts) based on a composite index that captured deprivation (25%), health (30%), education (15%) and infrastructure (30%) [49]. The aspirational districts program was aimed at reducing inter-district and inter-state disparity by bringing the state and the central governments together on various developmental schemes and promoting a healthy competition among the districts towards achievement of improved target outcomes. We also used a binary indicator for aspirational districts in our analysis.

In order to control for district specific heterogeneity in demographic and socio-economic indicators, we also used several related variables from the 2011 census of India that included rural population, literacy, gender, social and religious minority and population density as a percentage or proportion of district population in the census data.

We created a comprehensive district level data set using the variables from various data sources as described above. Table 1 in the appendix describes different variables that we used for our analysis and the secondary data source from which it was obtained. Multiple linear regression models were used for each vaccination phase with the proportion of population vaccinated with the first dose of vaccine during the given phase as the dependent variable. The phase wise regression tables are provided in table 7 and table 8 in section A.2 in the appendix. Columns P1-P4 in table 7 and 8 denote the vaccination phase whereas subscripts 1 and 2 for phase 4 denote the first and the second month of phase 4. The explanatory variables in table 7 regression included covid variables (i.e. caseload and death rates during each phase) and health infrastructure variables along with demographic and socio-economic variables. The regressions in table 8 also included indicators of health and education from the NFHS-5 data in addition to the explanatory variables used in table 7 regressions. We used state fixed effects to control for unobserved state specific effects. In addition, we also use robust standard errors clustered at the state level in order to account for error dependence within districts belonging to the same state. Best fit models reported in table 7 and 8 were selected based on adjusted R-squared values and measures of information criteria (ie. with lowest AICs and BICs).

## Results and Discussion

Covid vaccines in India were provided free of charge to all. Despite that people might not perceive it to be costless due to several opportunity costs and unobserved psychological costs, lack of trust and uncertainty surrounding vaccination, which can lead to vaccine hesitancy. Vaccine hesitancy can be a major driver of low demand and subsequently lower vaccination rates in spite of the availability of a vaccine. Therefore, availability of a vaccine does not mean that people will rush to get inoculated. The observed vaccination rates are the outcome of a complex interaction between the supply as well as the demand for the vaccine, and just one component alone.

Reported infection rates in a district, while likely to be indicative of actual infection rates, were also confounded with covid testing efforts. Districts that implemented more rigorous covid testing were also likely to report a higher number of covid cases compared to districts that had lower covid testing rates. On the contrary, unlike reported infection rates, covid fatalities were far more closely associated with the actual infection rate and were unlikely to be influenced by covid testing efforts. Both covid reported infection and death rates were likely to be correlated with vaccination rates, and hence we include these variables in our regression.

For all vaccination phases, we observed vaccination rates to be positively correlated with reported infection rates in the preceding vaccination phase. This might be driven by higher awareness about covid among people in districts with higher reported infection rates either due to higher testing efforts or prevalence of actual higher infection rates around them. We also included reported infection rate for the current phase, which stood out to be positive and significant only during the first month of phase 4. The initial first month of phase 4 coincided with the deadly second wave of covid infections in India. Therefore, it might be possible that the current infection rates were salient in driving the decision to get vaccinated. The association between covid deaths and vaccination rates were significant for phase 1 and phase 4 only. While the coefficient was negative for deaths as proportion of district population, it was positive for deaths as proportion of reported infection rate for the above period.

The above discussion tries to think of possible reasons for the observed relationship between past and current reported infection and deaths, which tell a demand side story. However, it cannot be undermined that covid infection and deaths would have been one of the critical inputs in designing vaccine allocation to different districts. Vaccination rates would have been lower in districts that were constrained due to limited supply of vaccines. Since, we had analysed the aggregate vaccination rates during an entire phase instead of daily or weekly vaccination, it might not be unreasonable to assume that vaccine supply would have been less volatile during an entire phase than over a week or on a particular day. Therefore, while not impossible, it is less likely that a district would have faced severe supply constraints due to short supply of vaccine during the entire vaccination phase. Nonetheless, we would not attribute lower vaccination rates to low demand only but rather a combination of both demand and supply side factors [45, 50].

Health infrastructure burden was captured by the proportion of population covered per health centre. A lower burden is indicative of better health infrastructure and is positively associated with vaccination rates across all vaccination phases and the coefficient was significant except phase 1. Since phase 1 targeted vaccination of health workers and frontline workers, it was not surprising to not find a significant correlation between health infrastructure and vaccination rates across districts.

Covid vaccination was available only at vaccination sites on demand which implied that individuals had to travel to the vaccination centres to get vaccinated. Health infrastructure density measures the average number of health centres per square kilometre in a district. While a lower density indicates greater distance from the health centre, we found inverse and significant correlation between health infrastructure density and covid vaccination rates in early phase 4. The beginning of phase 4 covid vaccination in India coincided with the deadly second wave (as can be seen in figure 1) with high covid infections and deaths along with multiple localised lockdowns, which adversely affected densely populated areas more severely. Given that population density and health infrastructure density are positively correlated, the above finding might not be surprising. Moreover, besides health centres, schools, community centres and other public places were also converted into vaccination centres. Therefore, the health infrastructure density variable might not be completely capturing the average distance to the vaccination centre as described.

With respect to different demographic and socio-economic variables, we did not find them to explain heterogeneity in the vaccination rates across districts during different vaccination phases except for phase 4. Vaccine attitudes are dynamic and evolve over time and space. Therefore, it was possible that these correlations could have emerged only during the fourth phase of vaccination, when India was hit by the deadly second wave of covid and the country witnessed the worst situation in the entire pandemic [51, 52, 53, 54, 55, 50, 56]. Vaccination rates were negatively correlated with higher proportion of rural population in a district but positively correlated with higher literacy rates. Covid vaccination campaigns in India required an online registration to get vaccinated during the period of our analysis. In addition, during the above period, the registration platform was only accessible in English. Therefore, appointment registration for covid vaccination required access to a smart phone and internet along with English literacy. These factors could be partly driving the relationship between literacy and vaccination. Population density too was positively correlated with higher vaccination rates. There is anecdotal evidence that suggested high vaccine hesitancy among the rural population during the beginning of the fourth phase, which aligns with the “co-incidence dragon” [Post hoc ergo propter hoc: after this, therefore because of this] in the literature [9, 57]. As stated earlier, roll out of covid vaccination in India prioritised the section of the population most susceptible to covid infection and fatalities. This section of the population included the elderly and those with comorbidities who had been vaccinated with the first dose of covid vaccine but yet to be vaccinated with the second dose of covid vaccine. It is important to mention that covid vaccines in India and elsewhere were mostly two shot vaccines. High efficiency would require that both doses of vaccines be administered in order to provide protection against fatal infection. During the second covid wave which coincided with the early fourth phase of vaccination, several deaths occurred among the vaccinated people, who were mostly vaccinated with the first dose of covid vaccine. Anecdotal evidence suggests that this led to the belief that vaccines were deadly and caused deaths giving way to coincidence dragon bias, particularly in the rural areas [58, 59, 60, 61]. Nonetheless, we cannot assert that coincidence dragon bias led to a reduction in vaccination numbers during the fourth phase. Given the severity of the second wave of covid, it cannot be ruled out that vaccine distribution would have been prioritised in favour of urban and more densely populated regions to contain the spread of infection which too could have adversely affected the vaccination drive in the rural areas.

There were no compelling correlations between social and religious groups that can explain the heterogeneity in vaccination rates consistently. During phase 4 of vaccination, districts with a higher proportion of SC population had lower vaccination rates. Vaccination rates were lower among the districts with relatively higher muslim population during phase 2. However, during the second month of phase 4, vaccination outcomes were relatively higher in districts with higher muslim populations. For muslims, Ramadan, the holy month of fasting, coincided with the first month of phase 4. Therefore, it could be the case that vaccination uptake was higher among muslims once Ramadan concluded [62, 63, 64].

We found vaccination rates to be correlated with selected maternal health indicators. Higher maternal protection against neonatal tetanus was positively correlated with covid vaccination rates. During phase 4, districts that had higher pregnancy rates between the age of 15 and 19 years and higher percentage of children exclusively breastfed had lower vaccination rates. Women of child bearing age were eligible for vaccination during the fourth phase of vaccination, but lack of scientific evidence and clear communication surrounding potential side-effects of the vaccine on a pregnant or a breastfeeding woman could have driven this finding [65, 66, 67].

With respect to child health indicators, districts with a higher proportion of wasted children had lower vaccination rates. We also found positive and significant correlation between complete immunisation of children and covid vaccination rates during phase 3 and 4 of the vaccination [68, 69]. However, for given complete immunisation levels, the correlation between polio vaccination and covid vaccination were negative. It is important to note that unlike all other vaccines in the complete immunisation schedule that are invasive, polio vaccines are oral vaccines. Hence, the negative correlation of covid vaccine with polio vaccine might be indicative of aversion towards invasive medical procedures that could have built some hesitancy around covid vaccine, which too was invasive. Moreover, adult vaccination is not common in India unlike child immunisation, which too could have created a resistance to get vaccinated [70].

A positive and significant correlation between covid vaccination rate and proportion of population on medication for blood pressure during phase 2 and phase 3 was observed. Since, blood pressure and hypertension was one of the identified co-morbidities associated with covid infection, covid vaccination rates could have been higher. We found some positive correlation between health insurance coverage and vaccination rates too. Vaccination rates were also higher in aspirational districts during the initial first two phases of vaccination.

We compiled district level data from various secondary sources at the district level that facilitated the warranted heterogeneity analysis. We would like to clearly state that the observed relationship in our analysis should not be interpreted as a causal relationship between vaccination rates and other explanatory variables. There might be some element of causality in our evidence, which would have been invaluable if it could be identified, a causal analysis is limited due to unavailability of requisite data. We also understand that there could be mismeasurement errors particularly with respect to covid variables due to significant underreporting, yet our analysis would be defensible unless there are systematic errors in measurement of variables. However, given all these limitations we make best efforts to bring the several pieces of data together to provide an exploratory analysis of covid vaccination in India.

## Conclusion

Our study is an attempt to explain the variation in the observed covid vaccination rates across districts in India during the first six months of the covid vaccination drive. Vaccination decision being a complex interplay between demand and supply, observed vaccination outcome could be indicating a short supply or demand or interaction of both is difficult to identify the causal factors that can explain spatio-temporal heterogeneity in vaccination rates. Therefore, our analysis provides exploratory evidence surrounding vaccination rates based on which we build thoughtful insights and suggest possible reasons that could have led to the observed empirical relationships.

We use data from the first six months of covid vaccination drive in India that we combine with several other administrative data to create a unique data set that facilitates heterogeneity analysis across different vaccination phases and districts. We find evidence of past reported infection rates positively correlated with higher first dose covid vaccination outcomes. Higher Deaths as a proportion of district population is associated with lower vaccination uptake but as a percentage of reported infection was positively correlated with first dose covid vaccination. Districts that on average had higher population burden per health centre also had lower covid vaccination rates. Districts with higher rural populations had lower vaccination rates whereas the association with literacy rate was positive. A higher vaccination among the population with higher blood pressure and hypertension (one of the comorbidities with covid infection) was observed while observed vaccination rates were lower among a higher proportion of pregnant women and breastfeeding mothers. Districts with higher percentage of children with complete immunisation were associated with higher covid vaccination rates whereas low vaccination rates were observed in districts that reported relatively higher percentage of wasted children.

Our research makes an important contribution to the area of vaccine research in the LMICs by using data from India that hosted the largest covid vaccination drive in the world. We uncover several associations between vaccination rates, health and demographic variables that provide insights, which can be used to design future research and investigations surrounding vaccinations in LMICs.

## Data Availability

All data produced in the present study are available upon reasonable request to the authors

## Appendix

### A.1. Summary Statistics

**Table 1:**
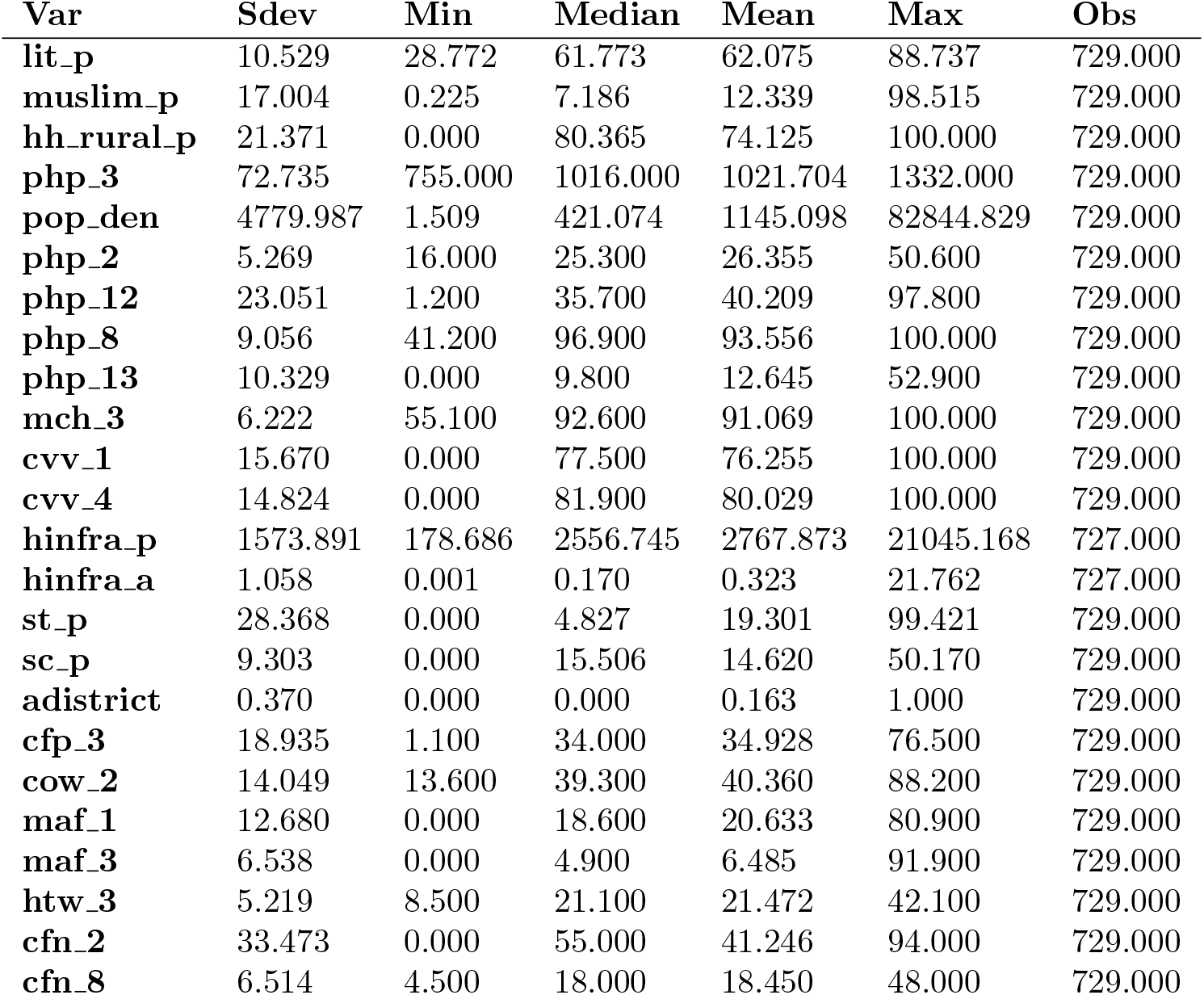
Demographic and health data variables

**Table 2:** Phase 1 Cowin data variables

| Var | Sdev | Min | Median | Mean | Max | Obs |
| --- | --- | --- | --- | --- | --- | --- |
| case_tp | 0.117 | -0.003 | 0.006 | 0.035 | 1.478 | 629.000 |
| case_cu | 0.676 | 0.000 | 0.440 | 0.639 | 6.045 | 629.000 |
| deth_cu | 0.011 | 0.000 | 0.004 | 0.008 | 0.111 | 629.000 |
| deth_case_cu | 0.839 | 0.000 | 1.052 | 1.184 | 4.988 | 625.000 |

**Table 3:** Phase 2 Cowin data variables

| Var | Sdev | Min | Median | Mean | Max | Obs |
| --- | --- | --- | --- | --- | --- | --- |
| case_tp | 0.131 | 0.000 | 0.009 | 0.054 | 1.475 | 629.000 |
| case_cu | 0.745 | 0.000 | 0.450 | 0.674 | 7.007 | 629.000 |
| deth_cu | 0.011 | 0.000 | 0.004 | 0.008 | 0.114 | 629.000 |
| deth_case_cu | 0.818 | 0.000 | 1.041 | 1.174 | 4.989 | 626.000 |

**Table 4:** Phase 3 Cowin data variables

| Var | Sdev | Min | Median | Mean | Max | Obs |
| --- | --- | --- | --- | --- | --- | --- |
| case_tp | 1.215 | 0.000 | 0.722 | 1.113 | 9.664 | 629.000 |
| case_cu | 0.816 | 0.000 | 0.477 | 0.728 | 7.356 | 629.000 |
| deth_cu | 0.012 | 0.000 | 0.004 | 0.008 | 0.116 | 629.000 |
| deth_case_cu | 0.728 | 0.000 | 1.026 | 1.112 | 4.270 | 626.000 |

**Table 5:** Phase 4.1 Cowin data variables

| **Var** | **Sdev** | **Min** | **Median** | **Mean** | **Max** | **Obs** |
| --- | --- | --- | --- | --- | --- | --- |
| **case_tp** | 0.693 | 0.000 | 0.421 | 0.642 | 8.183 | 629.000 |
| **case_cu** | 2.013 | 0.000 | 1.184 | 1.841 | 17.020 | 629.000 |
| **deth_cu** | 0.015 | 0.000 | 0.006 | 0.011 | 0.130 | 629.000 |
| **deth_case_cu** | 0.375 | 0.000 | 0.562 | 0.593 | 2.414 | 626.000 |

**Table 6:** Phase 4.2 Cowin data variables

| **Var** | **Sdev** | **Min** | **Median** | **Mean** | **Max** | **Obs** |
| --- | --- | --- | --- | --- | --- | --- |
| **case_tp** | 0.289 | 0.000 | 0.058 | 0.183 | 2.609 | 629.000 |
| **case_cu** | 2.560 | 0.000 | 1.700 | 2.483 | 20.299 | 629.000 |
| **deth_cu** | 0.023 | 0.000 | 0.010 | 0.019 | 0.159 | 629.000 |
| **deth_case_cu** | 0.490 | 0.000 | 0.722 | 0.789 | 3.768 | 626.000 |

### A.2. Regressions

**Table 7:** Regression table without the NFHS data variables

|  | *Dependent variable: pdos1* |  |  |  |  |
| --- | --- | --- | --- | --- | --- |
|  | (P1) | (P2) | (P3) | (P4<sub>1</sub>) | (P4<sub>2</sub>) |
| st_p | 0.01***<br>(0.001) |  |  |  |  |
| hh_rural_p |  |  |  | −0.03***<br>(0.01) | −0.03**<br>(0.01) |
| lit_p |  |  | 0.08***<br>(0.01) | 0.03***<br>(0.01) | 0.09***<br>(0.02) |
| php_3 |  | 0.002**<br>(0.001) | 0.003**<br>(0.001) |  |  |
| sc_p |  |  |  |  | −0.06**<br>(0.02) |
| muslim_p |  | −0.01**<br>(0.005) |  |  | 0.04***<br>(0.01) |
| pop_den |  |  | 0.0002***<br>(0.0001) |  | 0.001***<br>(0.0002) |
| hinfra_p |  | −0.0002***<br>(0.0000) | −0.0003***<br>(0.0001) | −0.0002***<br>(0.0001) | −0.001***<br>(0.0001) |
| hinfra_a |  |  |  |  | −4.69***<br>(1.03) |
| case_cu | 0.67***<br>(0.05) | 1.16***<br>(0.10) | 2.04***<br>(0.15) | 0.19***<br>(0.06) | 1.54***<br>(0.15) |
| deth_cu | −12.69***<br>(3.28) |  |  |  | −80.44***<br>(15.65) |
| deth_case_cu | 0.09**<br>(0.04) |  |  | −0.87***<br>(0.24) | 1.25***<br>(0.47) |
| case_tp |  |  |  | 0.81***<br>(0.20) |  |
| Constant | −0.03<br>(0.13) | −0.32<br>(1.03) | −5.83***<br>(1.70) | 4.36***<br>(0.99) | 2.54<br>(2.16) |
| State FE | Yes | Yes | Yes | Yes | Yes |
| Observations | 625 | 629 | 629 | 626 | 626 |
| AIC | 604.79 | 2030.32 | 2561.77 | 2279.99 | 3195.05 |
| BIC | 751.23 | 2190.3 | 2726.2 | 2439.81 | 3372.62 |
| R<sup>2</sup> | 0.64 | 0.70 | 0.78 | 0.66 | 0.72 |
| Adjusted R<sup>2</sup> | 0.62 | 0.68 | 0.77 | 0.65 | 0.71 |

**Table 8:** Regression table with the NFHS data variables

|  | *Dependent variable: pdos1* |  |  |  |  |
| --- | --- | --- | --- | --- | --- |
|  | (P1) | (P2) | (P3) | (P4<sub>1</sub>) | (P4<sub>2</sub>) |
| st_p | 0.01***<br>(0.001) |  | -0.01***<br>(0.005) |  |  |
| lit_p |  |  |  |  | 0.12***<br>(0.02) |
| females per 1,000 males |  |  |  |  | -0.01***<br>(0.002) |
| sc_p |  |  |  |  | -0.07***<br>(0.02) |
| muslim_p |  | -0.01***<br>(0.005) |  |  | 0.04***<br>(0.01) |
| adistrict | 0.15***<br>(0.05) | 0.31**<br>(0.14) |  |  |  |
| hh_rural_p |  |  |  | -0.04***<br>(0.01) |  |
| pop_den |  |  |  |  | 0.001***<br>(0.0002) |
| hinfra_p |  | -0.0002***<br>(0.0000) | -0.0003***<br>(0.0001) | -0.0002***<br>(0.0001) | -0.001***<br>(0.0001) |
| hinfra_a |  |  |  |  | -3.81***<br>(1.02) |
| case_cu | 0.61***<br>(0.05) | 1.09***<br>(0.10) | 2.03***<br>(0.15) | 0.34***<br>(0.09) | 1.53***<br>(0.14) |
| deth_cu | -11.16***<br>(3.20) |  |  | -26.52**<br>(10.58) | -72.47***<br>(15.42) |
| deth_case_cu | 0.08**<br>(0.04) | 0.02<br>(0.09) | 0.09<br>(0.16) | -0.38<br>(0.29) | 1.33***<br>(0.47) |
| Population ≤ 15 years | -0.02***<br>(0.01) | | | | |
| case_tp |  |  |  | 0.77***<br>(0.19) |  |
| HH with health insurance | 0.01***<br>(0.002) |  |  | 0.02**<br>(0.01) | 0.05***<br>(0.02) |
| Female sterilization | -0.01***<br>(0.002) |  |  |  |  |
|  | (P1) | (P2) | (P3) | (P4<sub>1</sub>) | (P4<sub>2</sub>) |
| HH improved drinkingwater source |  | 0.02**<br>(0.01) |  |  |  |
| Child attending preschool |  | -0.02***<br>(0.01) |  |  |  |
| Women schooling $\geq 10$ years | | | 0.05***<br>(0.01) | | |
| married before age 18 |  |  |  |  | 0.09***<br>(0.02) |
| age 15-19 pregnant |  |  |  |  | -0.13***<br>(0.05) |
| protected - neonatal tetanus |  |  |  |  | 0.05*<br>(0.03) |
| child vaccination |  |  | 0.05**<br>(0.02) |  | 0.08**<br>(0.04) |
| polio vaccination |  |  | -0.06**<br>(0.02) |  | -0.10**<br>(0.04) |
| f taking meds for bp |  | 0.07***<br>(0.01) | 0.10***<br>(0.02) |  |  |
| child exclusively breastfed |  |  |  | -0.01***<br>(0.002) |  |
| weight for height |  |  |  | -0.05***<br>(0.01) |  |
| Constant | 0.21<br>(0.29) | -1.72*<br>(0.98) | -1.10<br>(0.87) | 5.40***<br>(0.94) | -2.50<br>(3.96) |
| State FE | Yes | Yes | Yes | Yes | Yes |
| Observations | 625 | 626 | 626 | 626 | 626 |
| AIC | 571.81 | 1996.14 | 2527.83 | 2255.96 | 3174.98 |
| BIC | 736.01 | 2164.84 | 2696.52 | 2429.1 | 3379.19 |
| R<sup>2</sup> | 0.66 | 0.71 | 0.79 | 0.68 | 0.74 |
| Adjusted R<sup>2</sup> | 0.64 | 0.69 | 0.78 | 0.66 | 0.72 |

## Notes

### Competing Interest Statement

The authors have declared no competing interest.

### Funding Statement

This study did not receive any funding

